# Machine learning models for predicting medium-term heart failure prognosis: Discrimination and calibration analysis

**DOI:** 10.1101/2024.12.17.24319186

**Authors:** Takuya Nishino, Katsuhito Kato, Shuhei Tara, Daisuke Hayashi, Tomohisa Seki, Toru Takiguchi, Yoshiaki Kubota, Takeshi Yamamoto, Mitsunori Maruyama, Eitaro Kodani, Nobuaki Kobayashi, Akihiro Shirakabe, Toshiaki Otsuka, Shoji Yokobori, Yukihiro Kondo, Kuniya Asai

**Affiliations:** Department of Health Care Administration, Nippon Medical School, Tokyo, Japan; Department of Hygiene and Public Health, Nippon Medical School, Tokyo, Japan; Department of Cardiovascular Medicine, Nippon Medical School, Tokyo, Japan; Department of Pharmaceutical Service, Nippon Medical School Hospital, Tokyo, Japan; Department of Healthcare Information Management, The University of Tokyo Hospital, Tokyo, Japan; Department of Emergency and Critical Care Medicine, Nippon Medical School, Tokyo, Japan; Division of Cardiovascular Intensive Care, Nippon Medical School Hospital, Tokyo, Japan; Department of Cardiovascular Medicine, Nippon Medical School Musashi-Kosugi Hospital, Kawasaki, Japan; Department of Cardiovascular Medicine, Nippon Medical School Tama Nagayama Hospital, Tama, Japan; Department of Cardiovascular Medicine, Nippon Medical School Chiba Hokusoh Hospital, Inzai, Japan; Division of Intensive Care Unit, Nippon Medical School Chiba Hokusoh Hospital, Inzai, Japan; Department of Urology, Nippon Medical School, Tokyo, Japan

**Keywords:** mortality, rehospitalization, receiver operating characteristic curve, precision-recall curve, calibration slope, Brier score

## Abstract

**Background:** The number of patients with heart failure (HF) is increasing with an aging population, shifting care from hospitals to clinics. Predicting medium-term prognosis after discharge can improve clinical care and reduce readmissions; however, no established model has been evaluated with both discrimination and calibration.

**Objectives:** This study aimed to develop and assess the feasibility of machine learning (ML) models in predicting the medium-term prognosis of patients with HF.

**Methods:** This study included 4,904 patients with HF admitted to four affiliated hospitals at Nippon Medical School (2018–2023). Four ML models—logistic regression, random forests, extreme gradient boosting, and light gradient boosting—were developed to predict the endpoints of death or emergency hospitalization within 180 days of discharge. The patients were randomly divided into training and validation sets (8:2), and the ML models were trained on the training dataset and evaluated using the validation dataset.

**Results:** All models demonstrated acceptable performance as assessed by the area under the precision-recall curve. The models showed favorable agreement between the predicted and observed outcomes in the calibration evaluations with the calibration slope and Brier score. Successful risk stratification of medium-term outcomes was achieved for individual patients with HF. The SHapley Additive exPlanations algorithm identified nursing care needs as a significant predictor alongside established laboratory values for HF prognosis.

**Conclusions:** ML models effectively predict the 180-day prognosis of patients with HF, and the influence of nursing care needs underscores the importance of multidisciplinary collaboration in HF care.

**Clinical Trial Registration:** URL: https://www.umin.ac.jp/ctr; unique identifier: UMIN000054854

## Introduction

Heart failure (HF) is a global health challenge, affecting an estimated 64.3 million people worldwide [1], with a growing number of hospitalizations [2,3]. This situation has raised concerns about strained healthcare provision and a marked increase in medical costs, leading to a shift in patient care for HF from hospitals to clinics [4]. In this context, effective cooperation between hospitals and clinics is essential. Routine patient care is delegated to primary care physicians in clinics, while hospital doctors conduct regular specialized checkups. Therefore, predicting the risk of medium-term deterioration after hospital discharge can assist primary care physicians in managing these patients. Furthermore, sharing this prediction among multiple healthcare professionals contributes to planning preventive care tailored to individual patients, thereby reducing the risk of readmission in patients with HF.

Machine learning (ML) models developed by integrating multiple factors have been applied to predict outcomes in patients with HF and are expected to outperform conventional statistical methods in predictive accuracy [5]. Although many studies have demonstrated the utility of ML models in predicting short-term outcomes, such as morbidity, in-hospital mortality, and 30-day rehospitalization rates [6–8], the effectiveness of these models in predicting medium- to long-term prognosis remains unclear. Previous ML models have predominantly relied on pathophysiological factors of the disease, such as clinical laboratory values, electrocardiographic features, echocardiographic findings, and medications [9,10]. However, these models have not accounted for information recorded by medical professionals other than doctors, such as physical conditions, nursing care needs, medication adherence, and the social background of patients [2,11]. Comprehensive integration of these variables is crucial for improving medium-term prognostic accuracy.

This study aimed to develop and validate ML models that incorporate the physical status of patients in addition to clinical laboratory data and treatment details to predict the prognosis of patients with HF within 180 days of discharge. We selected ML models as conventional logistic regression models using linear feature extraction and tree-based prediction models using nonlinear feature extraction. We evaluated the discrimination of these models using the area under the receiver operating characteristic curve (AUROC), the area under the precision-recall curve (AUPRC), and the agreement between the models’ predicted probabilities and observed outcomes by calibration.

## Methods

### Study Design and Data Collection

This multicenter cohort study was conducted using an inpatient database from four affiliated hospitals of Nippon Medical School, including Nippon Medical School Hospital, Musashi Kosugi Hospital, Tama Nagayama Hospital, and Chiba Hokusoh Hospital. The database was constructed using Diagnosis Procedure Combination (DPC) data and laboratory test values from medical records. Data for this study were accessed on July 21, 2024. The authors had no access to information that could identify individual participants during or after data collection. This study was approved by the Central Ethics Review Committee of Nippon Medical School (approval number M-2024-178) and conducted in accordance with the Declaration of Helsinki. Participant consent was obtained using an opt-out method. Reporting followed the Transparent Reporting of a Multivariable Prediction Model for Individual Prognosis or Diagnosis (TRIPOD) guidelines [12].

### Patient Selection and Endpoints

Patients included in this study were those hospitalized between April 2018 and September 2023, aged ≥18 years, and diagnosed with HF. HF was defined as having a brain natriuretic peptide (BNP) level of ≥100 pg/mL or an N-terminal pro-BNP (NT-proBNP) level ≥300 pg/mL during hospitalization, in alignment with international definitions of HF [13]. Exclusions were made for patients with a hospital stay of <5 days, those discharged due to death or transfer to another hospital, or those lacking event occurrence with <180 days of follow-up. The endpoint was a composite of all-cause mortality and emergency readmission within 180 days of discharge.

### Variables

The DPC database captured data on all hospitalized patients, including demographics (age, sex), physical metrics (height, weight, body mass index [BMI], calculated by dividing body weight [kg] by the square of height [m^2^]), and clinical details (prior emergency hospitalization, comorbidities and procedures during hospitalization, and medications at discharge) [14]. The number of prior emergency hospitalizations was defined as those within 180 days after discharge. The medications at discharge recommended in the guidelines for HF and coronary artery disease were defined as those of guideline-directed medical therapy (GDMT) (**Table S1**) [15,16].

Medications other than those of GDMT were defined as those not included in the GDMT (ni-GDMT). In this study, we focused on the number of ni-GDMT medications because they are considered to reflect the number of comorbidities, their severity, and polypharmacy. The patient’s physical status was evaluated using the nursing care needs score at discharge [14]. This score is the sum of the level of assistance required for turning over, transferring, oral hygiene, eating, and changing clothes (0 points: no assistance, 1 point: partial assistance, 2 points: full assistance), whether medical instructions were understood (0 points: yes, 1 point: no), and the presence of risky behavior (0 points: no, 2 points: yes) (**Table S2**).

Blood test data at admission and discharge were obtained from medical records and defined as the first sampling within 3 days of admission and the last sampling within 14 days before discharge, respectively. After applying the inclusion criteria, BNP was converted to NT-proBNP using the following conversion formula: NT-proBNP = 10^(1.1 × log10[BNP] + 0.57)^ [17]. Numerical data were scaled using the Standard Scaler.

### Imputation of Missing Data

Missing data for BMI and blood test findings were imputed using a single imputation implemented in the Python MissForest package [18,19]. All variables were used for analysis because the missing rate for all variables was <20%, which is the threshold for exclusion.

### Predictive Model Development

We randomly divided the dataset into 80% training and 20% test sets using stratified sampling to preserve the endpoint occurrence rates of the original population. We used the following ML algorithms as predictive models: conventional logistic regression (LR), tree-based random forest (RF), extreme gradient boosting (XGB), and light gradient boosting machine (LGBM) [20–22].

In this study, we used recursive feature elimination with cross-validation (RFECV) of each model to select the optimal subset of features specific to each model. RFECV is a robust method that recursively eliminates less important features and builds a model with the remaining features to identify the optimal subset. Specifically, RFECV was implemented for each model using a 10-fold stratified cross-validation strategy to maximize the AUROC. After selecting the most relevant features, hyperparameter tuning was performed for each model using a grid search with cross-validation to determine the optimal model parameters. This step involved evaluating various combinations of hyperparameters and ultimately selecting the combination that maximized the AUROC for each model (**Table S3**). As a result of the optimization process (**Table S4**), the hyperparameter set with the highest AUROC score for each model was identified. The models were calibrated using isotonic regression [23].

SHapley Additive exPlanations (SHAP) values were used to explain the output of the ML models [24]. SHAP values quantify the contribution of each feature to the predictions made by the models, allowing for a better understanding of the factors driving the model’s decisions.

### Model Validation

The bootstrap method was applied to the test set to evaluate the performance of the predictive models. A total of 2,500 bootstrap resamples were used to calculate 95% confidence intervals (CIs) for each performance metric. The discriminatory ability of the model was assessed using the AUROC and AUPRC [25,26]. For the calibration analysis, the predicted probabilities were divided into ten percentiles, and the mean predicted probability of the outcome and observed probability of the outcome for each bin were plotted. Two indices were calculated to evaluate the calibration: the calibration slope, which indicates the agreement between the predicted probabilities and observations, and the Brier score, which measures the accuracy of the probability predictions. A calibration slope closer to 1 and a Brier score closer to 0 indicate ideal model performance.

### Risk Classification

We classified the probabilities predicted by the ML models into three categories for risk stratification: Given that the probability of rehospitalization within 1 year for patients with HF is approximately 30% [27,28], the categories were defined as low risk (< 0.15), middle risk (>0.15 and <0.30), and high risk (≥0.30).

Survival analysis was performed using the Kaplan–Meier method, and the log-rank test was used to compare survival distributions between the stratified groups.

### Package for Analysis

All statistical analyses were performed using Python version 3.9.0 (Python Software Foundation, www.python.org) and R software version 4.2.2 Patched (R Foundation for Statistical Computing, Vienna, Austria). A two-tailed test was performed, and a P-value of <0.05 was considered statistically significant.

## Results

### Study Population and Baseline Characteristics

Among the 9,519 patients, we excluded 905 who were discharged because of death, 1,402 who were transferred to other facilities, 466 with a hospital stay <5 days, and 1,842 without events and a follow-up period <180 days. Therefore, 4,904 participants were included in the final analysis, with 3,923 (80%) patients allocated to the training dataset and 981 (20%) allocated to the validation dataset (**Fig 1**).

**Fig 1.**
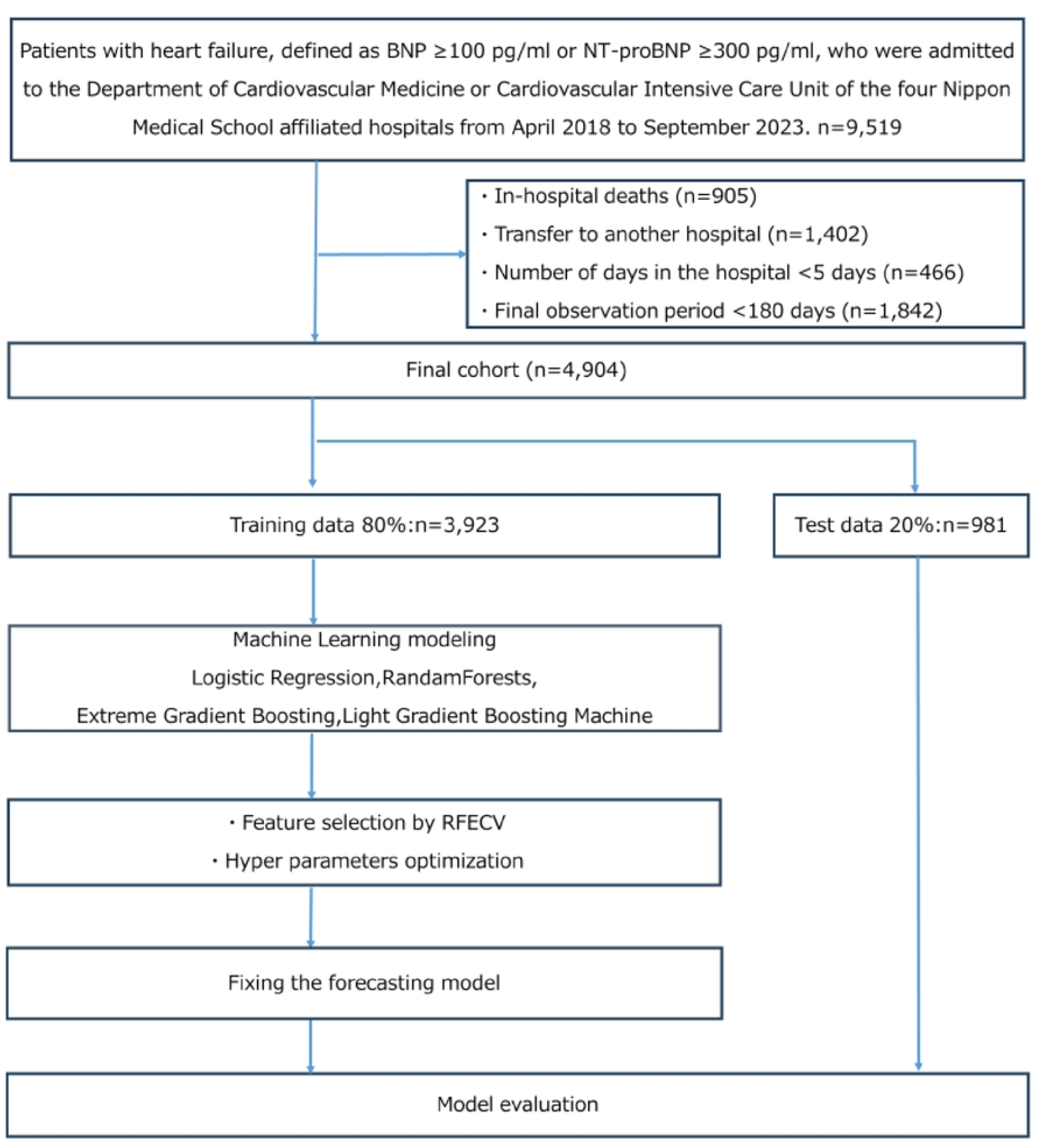
Flowchart for patient assignment and model development. BNP, brain natriuretic peptide; NT-proBNP, N-terminal pro-brain natriuretic peptide; RFECV, recursive feature elimination with cross-validation

**Table 1** shows the descriptive statistics of the variables included in the training and test datasets before data processing and the selected features. Outcomes occurred in 1,291 (26.3%) patients, and the features of the training and test datasets were well balanced. For the development of the ML models, out of a total of 61 variables, 28 features were selected for LR, 51 for RF, 52 for XGB, and 36 for LGBM (**Table S5**).

**Table 1.**
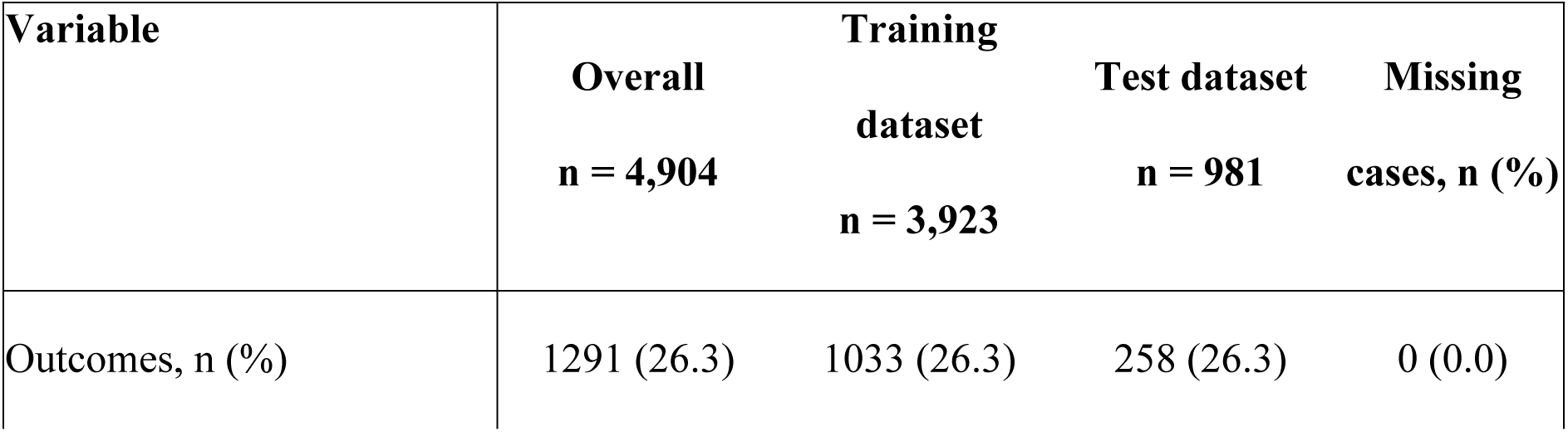
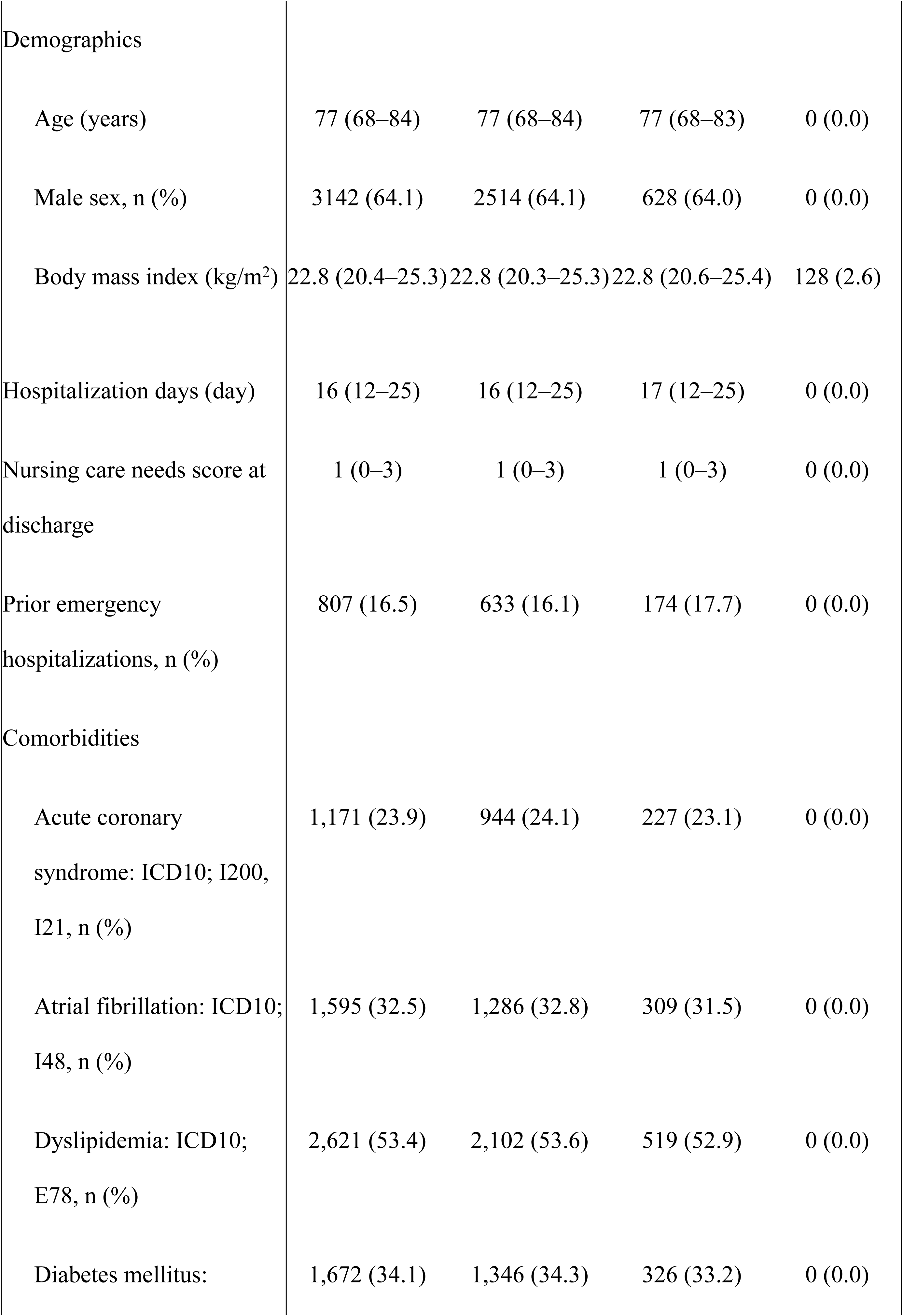
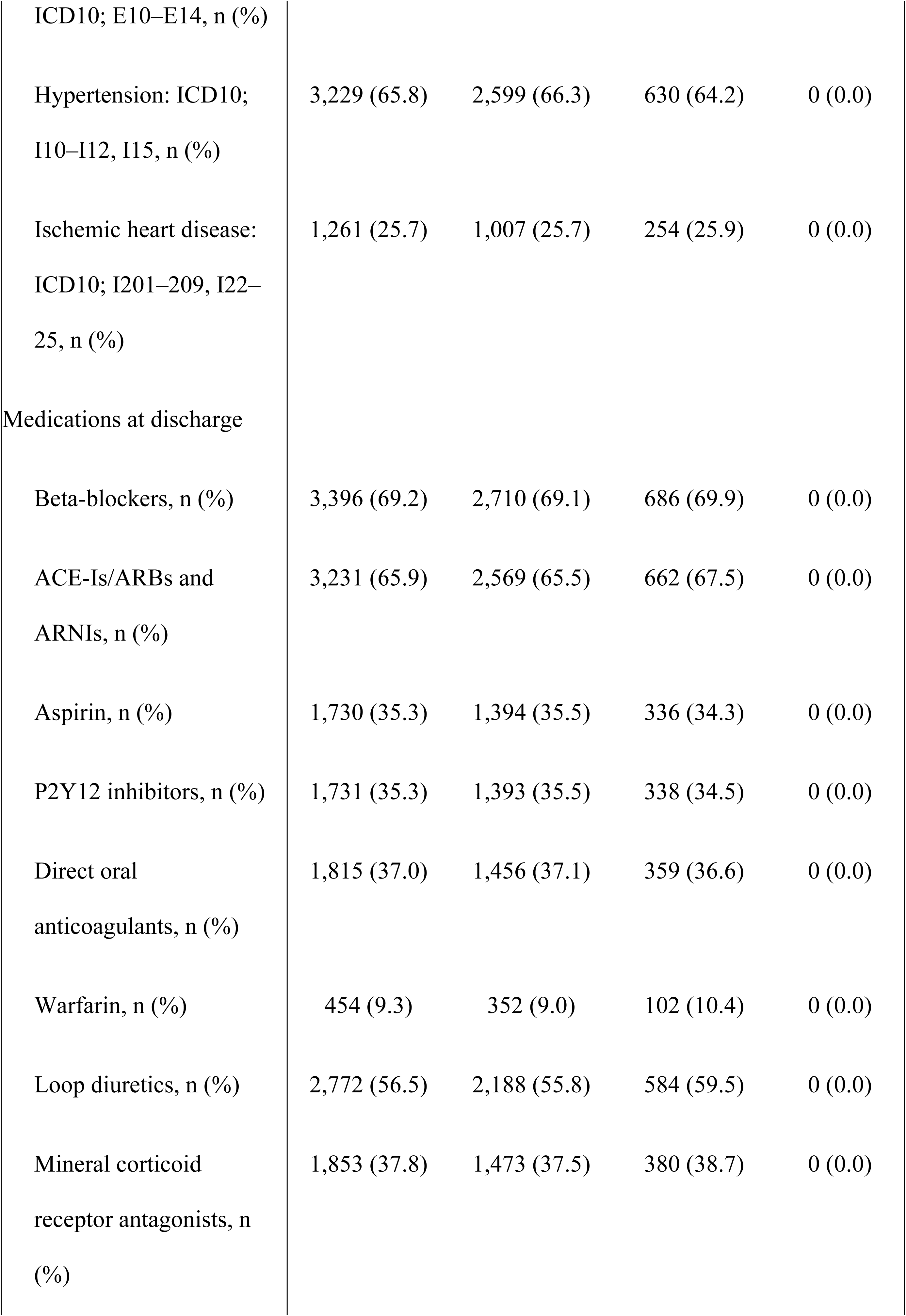
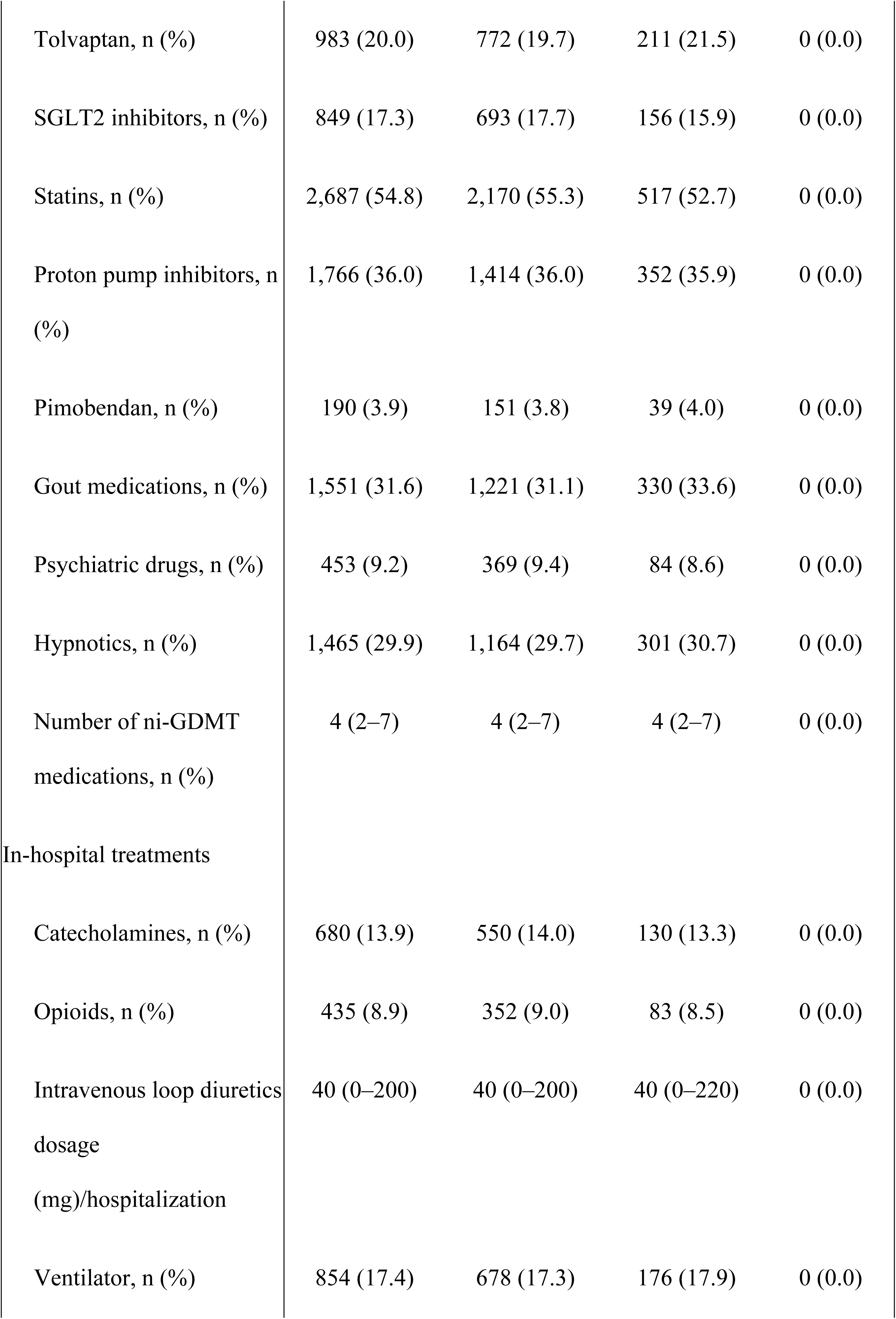
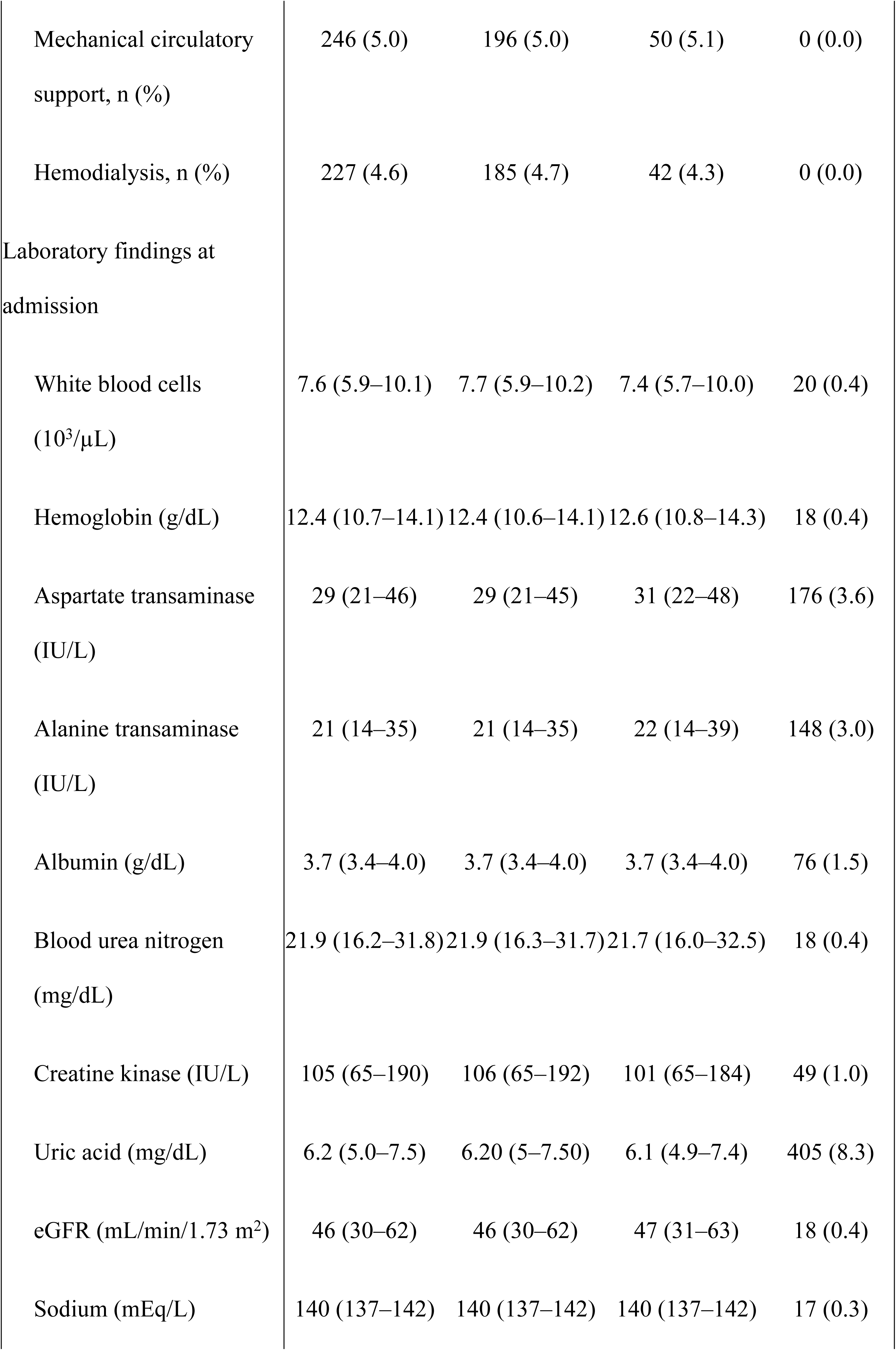
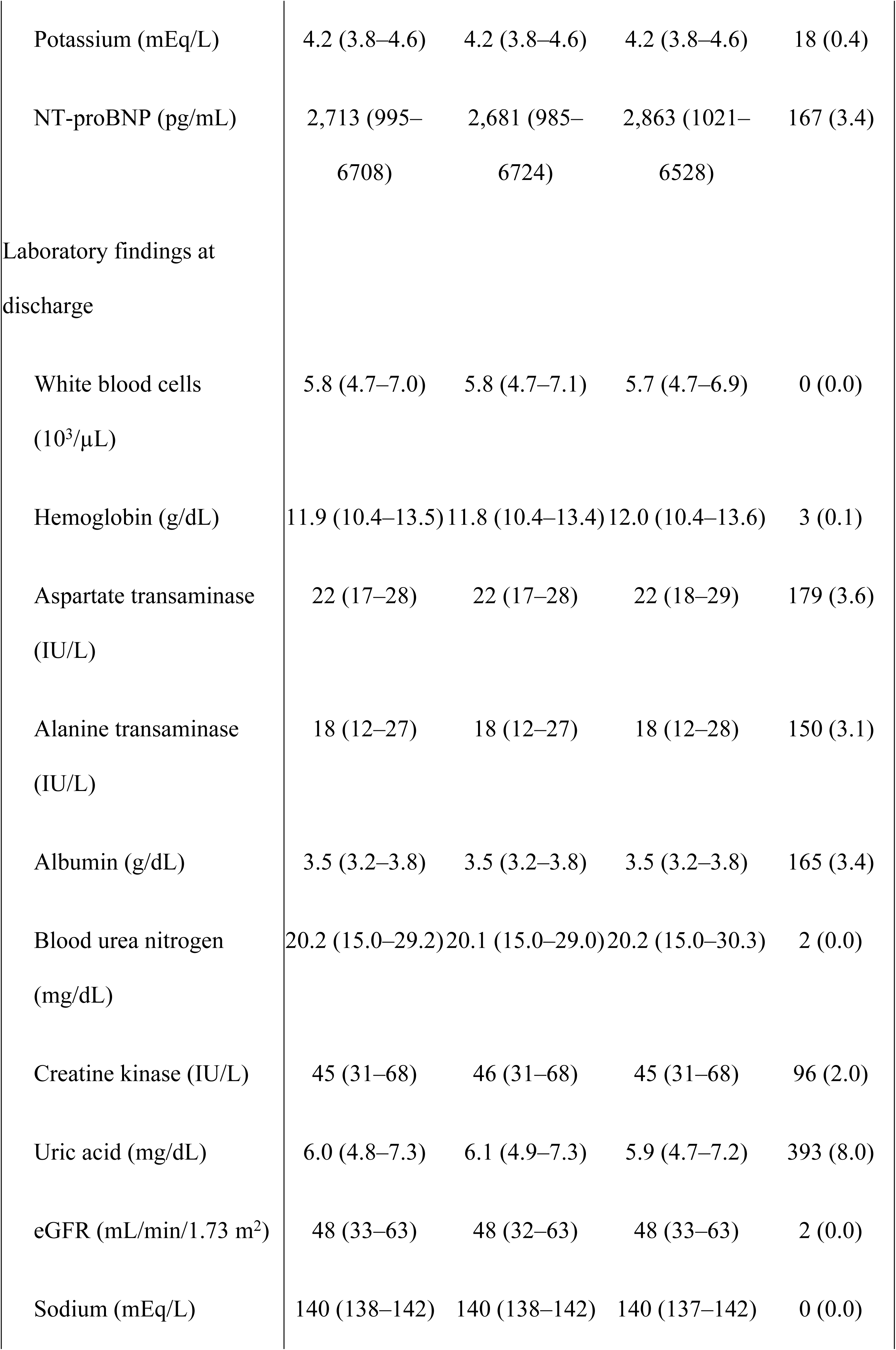
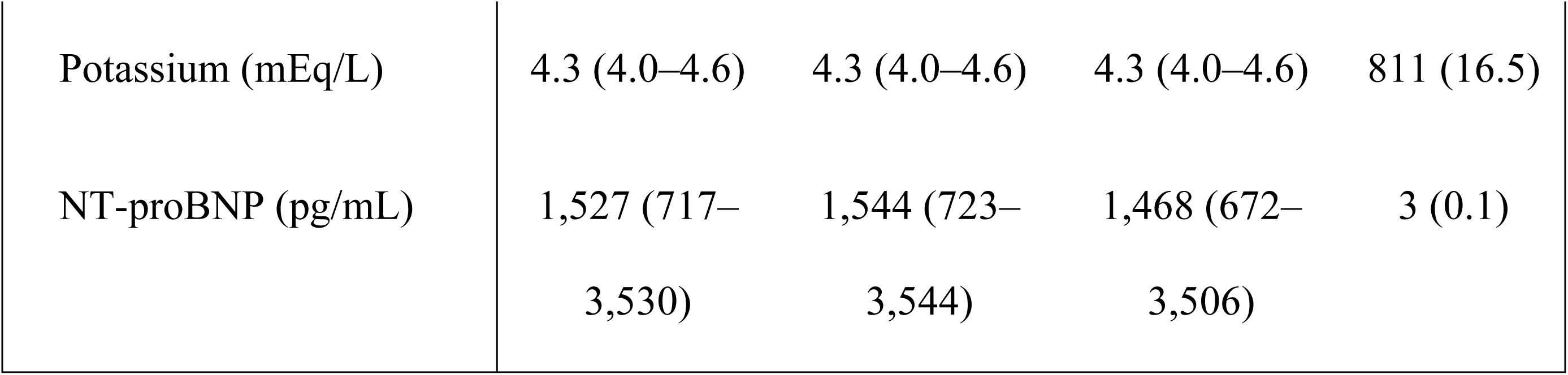
Patient characteristics.

Data are expressed as medians (interquartile ranges). ICD, International Statistical Classification of Diseases and Related Health Problems; ACE-Is, angiotensin-converting enzyme inhibitors; ARBs, angiotensin II receptor blockers; ARNIs, angiotensin receptor neprilysin inhibitors; SGLT2, sodium-glucose cotransporter 2; ni-GDMT, not included in guideline-directed medical therapy; eGFR, estimated glomerular filtration rate; NT-proBNP, N-terminal pro-brain natriuretic peptide

### Evaluation of Prediction Models

After training the models, the AUROC was calculated based on the results of the predicted events in the test dataset using the trained ML models. The AUROC values for LR, RF, XGB, and LGBM were 0.734 (95% CI, 0.698–0.769), 0.753 (95% CI, 0.718–0.786), 0.744 (95% CI, 0.708–0.778), and 0.752 (95% CI, 0.716–0.785), respectively (**Fig 2A**). Consequently, the models were evaluated by AUPRC, which provides more informative insights into binary classification predictive models with imbalanced data, and the values for LR, RF, XGB, and LGBM were 0.505 (95% CI, 0.445–0.568), 0.513 (95% CI, 0.454–0.579), 0.524 (95% CI, 0.463–0.589), and 0.515 (95% CI, 0.454–0.582), respectively (**Fig 2B**).

**Fig 2.**
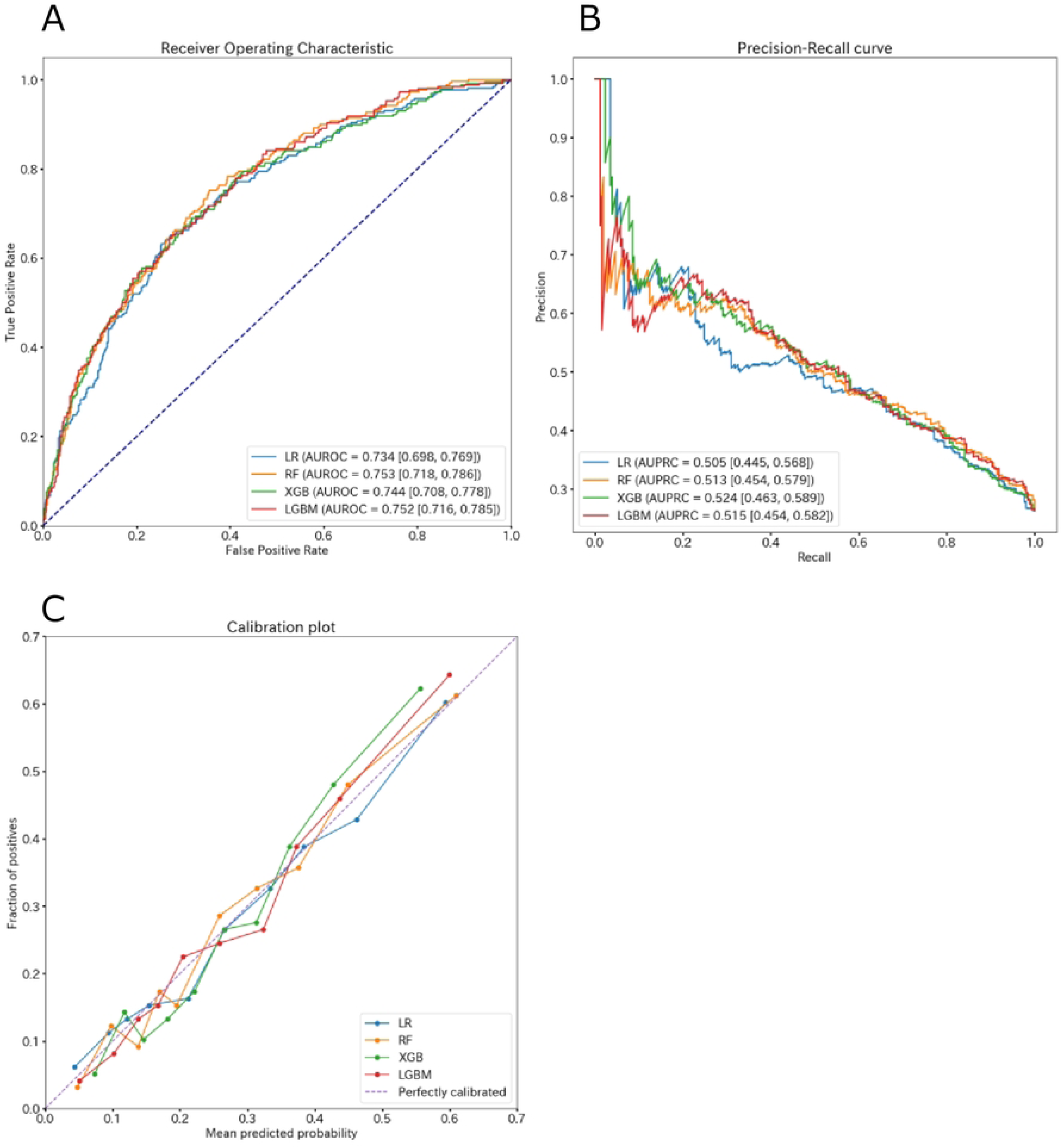
Model validations. (**A**) The receiver operating characteristics curve and area under the receiver operating characteristic (AUROC) curve of the models. AUROC is presented with a 95% confidence interval (CI). (**B**) The precision-recall curves and area under the precision-recall (AUPRC) curve of the models. The AUPRC is presented with a 95% CI. (**C**) Calibration plot of the models. LR, logistic regression; RF, random forests; XGB, extreme gradient boosting; LGBM, light gradient boosting machine

Calibration was assessed using a plot diagram of predicted probabilities and observations across the ten deciles of predicted risk, and favorable agreement between both values was observed in all models (**Fig 2C**). As calibration indexes for the models, values for the calibration slope (LR, 0.964; RF, 1.046; XGB, 1.197; LGBM, 1.088) and Brier score (LR, 0.167; RF, 0.164; XGB, 0.165; LGBM, 0.164) were calculated (**Table 2**). Furthermore, the observed incidence of events was compared across the three defined risk categories (low, middle, and high risk). The observed event rates for each stratified group were within the predicted probability ranges (**Table 3**). Survival analysis using the Kaplan–Meier method demonstrated that the cumulative observed event rate significantly increased with risk stratification across the four models (**Fig 3**).

**Fig 3.**
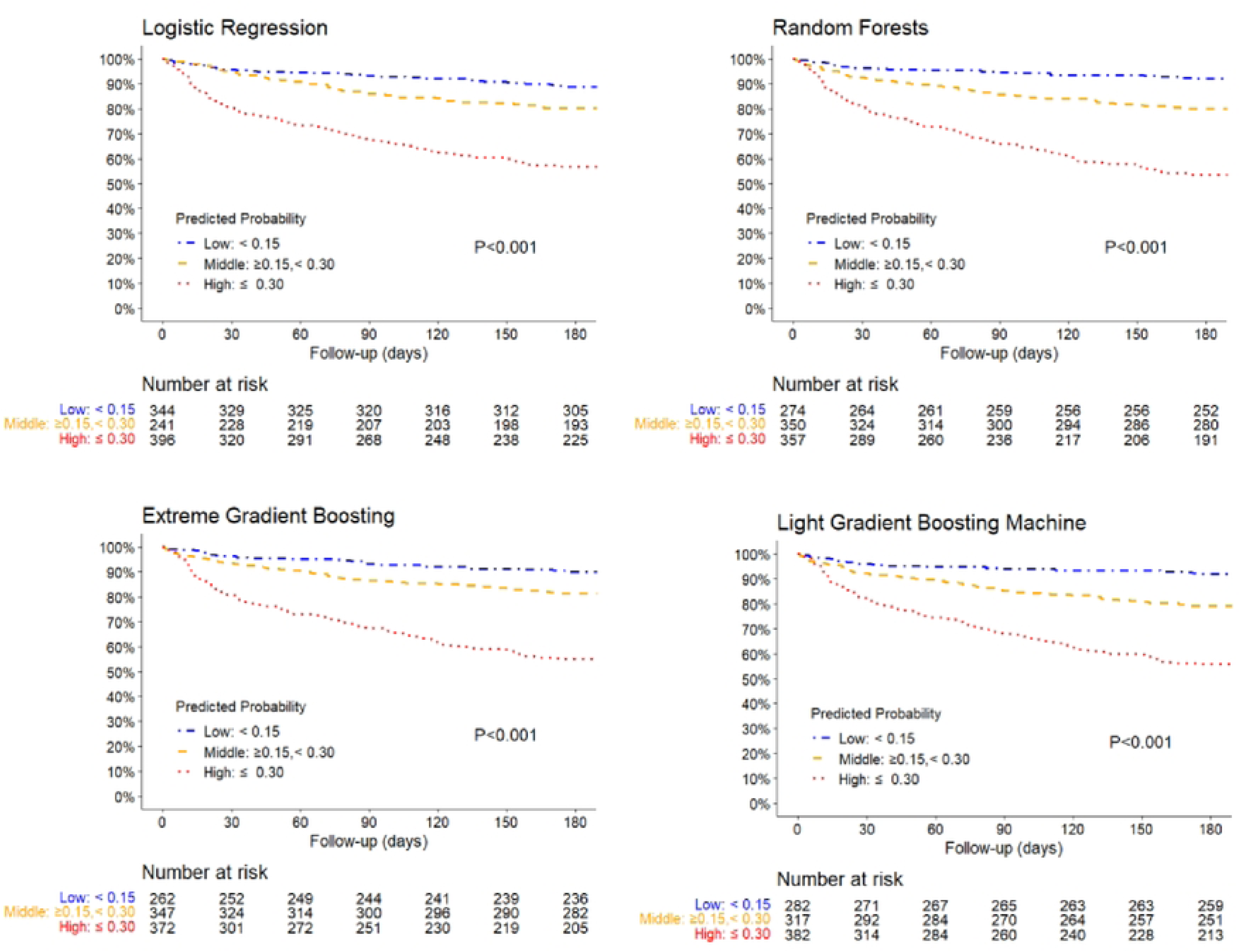
Kaplan–Meier curve analysis of 180-day all-cause mortality and emergency readmission. The cumulative incidence of events observed in each model is compared across the three risk categories (low, middle, and high risk).

**Table 2.** Calibration indexes.

| Model | Calibration slope (95% confidence interval) | Brier score (95% confidence interval) |
| --- | --- | --- |
| Logistic regression | 0.964 (0.804–1.120) | 0.167 (0.154–0.181) |
| Random forests | 1.046 (0.890–1.260) | 0.164 (0.151–1.177) |
| Extreme gradient<br>boosting | 1.197 (1.013–1.377) | 0.165 (0.153–0.178) |
| Light gradient<br>boosting machine | 1.088 (0.927–1.250) | 0.164 (0.152–0.177) |

**Table 3.** Observed outcomes for each risk stratum classified by predicted probability.

| Risk stratum<br>(range of predictive<br>probability) | Number of<br>patients<br>(total) | Number of<br>patients<br>(outcomes) | Observed<br>outcome rate |
| --- | --- | --- | --- |
| Logistic regression |  |  |  |
| Low: < 0.15 | 344 | 39 | 0.11 |
| Middle: $\geq 0.15$, < 0.30 | 241 | 48 | 0.20 |
| High: $\geq 0.30$ | 396 | 171 | 0.43 |
| Random forests |  |  |  |
| Low: < 0.15 | 274 | 22 | 0.08 |
| Middle: $\geq 0.15$, < 0.30 | 350 | 70 | 0.20 |
| High: $\geq 0.30$ | 357 | 166 | 0.46 |
| Extreme gradient boosting |  |  |  |
| Low: < 0.15 | 262 | 26 | 0.10 |
| Middle: $\geq 0.15$, < 0.30 | 347 | 65 | 0.19 |
| High: $\geq 0.30$ | 372 | 167 | 0.45 |
| Light gradient boosting machine |  |  |  |
| Low: $< 0.15$ | 282 | 23 | 0.08 |
| Middle: $\geq 0.15, < 0.30$ | 317 | 66 | 0.21 |
| High: $\geq 0.30$ | 382 | 169 | 0.44 |

### Model Interpretation

The contribution of each feature to the prediction of outcomes was visualized using the SHAP algorithm. The features were ranked in descending order of importance scores, with red indicating high influence and blue indicating low influence. Established risk factors for HF, such as age, estimated glomerular filtration rate (eGFR), NT-proBNP levels, hemoglobin levels, albumin levels, and the number of previous hospitalizations, were ranked among the top contributors in all four models. Furthermore, the number of ni-GDMT medications and nursing care needs scores, which were the features we focused on in this study, were identified as highly influential factors for outcomes (**Fig 4**).

**Fig 4.**
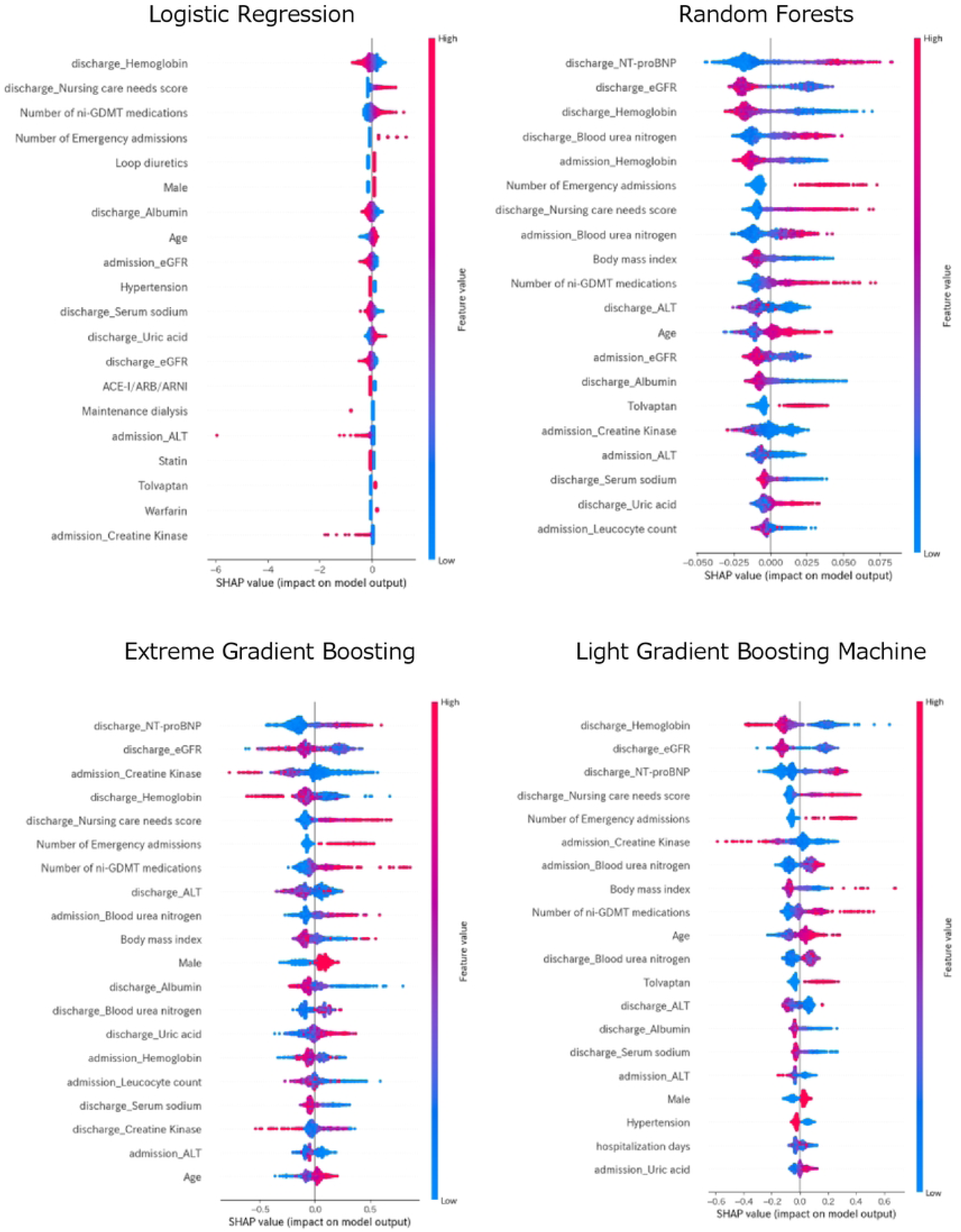
SHapley Additive exPlanations (SHAP) values in the models. The top 20 variables of SHAP analysis in each model are shown, along with the impact of their contribution to the prediction. eGFR, estimated glomerular filtration rate; ni-GDMT, not included in the guideline-directed medical therapy; ACE-I, angiotensin-converting enzyme inhibitor; ARB, angiotensin II receptor blocker; ARNI, angiotensin receptor neprilysin inhibitor; ALT, alanine aminotransferase; NT-proBNP, N-terminal pro-brain natriuretic peptide; AST, aspartate aminotransferase

## Discussion

In this multicenter retrospective cohort study, we developed ML models to predict all-cause mortality and emergency admission within 180 days of discharge in patients with HF. The models were trained on the training dataset and evaluated on the internal validation dataset. The discriminative power of each model fell within the range of AUROC (0.65–0.78) reported in previous studies on long-term prognosis prediction of patients with HF [6]. Moreover, the conventional LR model and tree-based models (RF, XGB, and LGBM) demonstrated favorable agreement between predicted and observed values in the calibration, and successful risk stratification was achieved in these models.

Discrimination refers to the ability of a predictive model to distinguish correctly between positive and negative cases. In this study, the tree-based ML models demonstrated equal or better discriminatory power than the conventional LR model did for both AUROC and AUPRC. This is consistent with the concept that tree-based approaches that make predictions based on nonlinear relationships are more suitable than LR assuming linearity, especially because some features, such as eGFR and BMI, follow a U-shaped curve [29,30].

The ML models in this study demonstrated favorable agreement in calibration, which is an indicator of how closely the model’s predicted probability matched the observed outcome. Although this evaluation method with calibration is important for stratifying the risk of individual patients and applying predictive models to clinical practice [31,32], there is a lack of model evaluation with calibration in a predictive model for HF [10,33]. In this study, the observed event rates in each stratum fell within the predicted probability ranges (low, middle, and high risk), suggesting that appropriate risk stratification for medium-term prognosis was achieved at the time of hospital discharge. In assessing the utility of a model, improperly structured calibration can be problematic because it leads to either over- or underestimation of risk [34]. Among the various algorithms reported as calibration methods for predicting models [35], isotonic regression, which is one of the oldest and most commonly used methods, was applied in this study [23]. This approach ensured reasonable calibration was achieved, as indicated by the calibration plot, and effectively addressed the challenge of over- and underestimation of risk. However, emerging research being conducted on more precise calibration algorithms that utilize neural networks suggests the potential for improvement by applying these methods [36].

The nursing care needs score and the number of ni-GDMT medications highly influenced the prediction across all models, as shown by the SHAP analysis, in addition to well-known prognostic factors of HF [37–39]. This indicates that daily physical activity, comorbidity status, and polypharmacy are crucial in the prognosis of patients with HF. However, only the nursing care needs score was available to characterize physical activity in this study, as our retrospectively collected dataset did not include other variables related to the patient’s physical condition. Frailty and comorbidity burden are associated with rehospitalization within 6 months of discharge in older patients with HF [40]. Moreover, based on the recent knowledge of multiple comorbidities or social determinants of health, several physical activity parameters and social factors should be added to the features of ML models [41,42].

We constructed the dataset for this study using DPC and clinical laboratory data that are automatically stored in an electronic format and can be readily exported. The use of DPC and electronic medical records has been standardized across Japanese university hospitals, and several clinical studies have utilized similar databases [43]. Therefore, our results are generalizable and can be easily confirmed using data from other hospitals for external validation. However, our dataset only included inpatients at a university hospital providing advanced and specialized treatments and did not reflect comprehensive HF care in the community region with hospital–clinic cooperation. Incorporating clinical data from primary care physicians in clinics and patient information (such as social background, physical activity, and living conditions) recorded by healthcare professionals other than doctors into the database would lead to the construction of a more accurate model for predicting the progression of HF. Building such a comprehensive database with a regional hospital–clinic– care team is the next challenge.

### Limitations

This study has some limitations. First, as a retrospective study, the explanatory variables for the ML models were limited to readily available clinical data with few missing values. Data such as echocardiographic and electrocardiographic findings, vital signs, and social factors were not included in this study. Second, this study was conducted solely in healthcare facilities in Japan; therefore, the findings may not apply to patients in other regions. Future studies should validate the ML models using independent external data from various regions and hospitals. Third, only four ML algorithms were used. Other ML algorithms, such as neural networks and support vector machines, may exhibit better predictive performances.

Finally, NT-proBNP levels ≥300 pg/mL and BNP levels ≥100 pg/mL were used to diagnose HF. However, data on the conversion between BNP and NT-proBNP suggest that the BNP level equivalent to an NT-proBNP level of 300 pg/mL is slightly lower, at approximately 60– 75 pg/mL [44]. This discrepancy might have led to the inaccurate inclusion of patients with HF. Additionally, because a formula for conversion between BNP and NT-proBNP was used for model development, patients assessed with BNP may not have been accurately evaluated.

## Conclusions

For patients with HF, we developed ML models to predict all-cause mortality and emergency admission in the medium term within 180 days after hospital discharge. This study demonstrates that ML models have favorable discrimination and agreement between predicted probabilities and observations. These ML models can be useful in reducing rehospitalization after discharge through risk stratification in individual patients. Furthermore, the influence of nursing care needs on prediction indicates the importance of multidisciplinary collaboration in HF care.

## Data Availability

The deidentified participant data will not be shared.

## Acknowledgements

The authors extend their sincere thanks to all the people involved in patient care, including emergency staff, technicians, medical engineers, nurses, pharmacists, physicians, and surgeons at Nippon Medical School Hospital, Musashi-Kosugi Hospital, Tama Nagayama Hospital, Chiba Hokusoh Hospital, and Nippon Medical School.

## Author contributions

Takuya Nishino: Data Curation, Formal analysis, Software, Writing-Original draft preparation. Katsuhito Kato: Conceptualization, Methodology, Data Curation, Writing-Original draft preparation. Shuhei Tara: Conceptualization, Data curation, Writing - Review & Editing, Project administration. Daisuke Hayashi: Conceptualization, Methodology, Writing - Review & Editing. Tomohisa Seki: Supervision, Validation, Visualization, Writing - Review & Editing. Toru Takiguchi: Validation, Visualization, Writing - Review & Editing. Yoshiaki Kubota: Conceptualization, Methodology, Resources. Takeshi Yamamoto: Data curation. Mitsunori Maruyama: Data curation. Eitaro Kodani: Data curation. Nobuaki Kobayashi: Data curation. Akihiro Shirakabe: Visualization, Data curation. Toshiaki Otsuka: Conceptualization, Supervision. Shoji Yokobori: Supervision. Yukihiro Kondo: Supervision. Kuniya Asai: Supervision, Funding acquisition, Project administration.

## Data statement

Data supporting the findings of this study are available from the corresponding author upon request.

